# Rethinking conceptualizations of adult ADHD and how care is provided: a qualitative study

**DOI:** 10.1101/2023.10.12.23296967

**Authors:** Callie M. Ginapp, Norman R. Greenberg, Grace Macdonald-Gagnon, Gustavo A. Angarita, Krysten W. Bold, Marc N. Potenza

## Abstract

**Background:** Attention-deficit/hyperactivity disorder (ADHD) is a common condition that frequently persists into adulthood. Adults with ADHD often have unmet needs including experiencing high rates of substance use disorders, incarceration, and unemployment. Despite having unmet needs, there is little research on how adults with ADHD conceptualize their condition and how they believe the care they receive could be improved. We aimed to better understand what adults with ADHD would like the medical community to know about what it is like to live with ADHD and to characterize possible misconceptions of ADHD they would like to see addressed.

**Method:** Nine online focus groups involving young adults (aged 18-35 years, N = 43) recruited from online communities for people with ADHD were conducted. Data were analyzed using an interpretive phenomenological analysis framework.

**Results:** Individually, participants reported wanting increased support, education, and rejection of stigma. Structurally, participants reported desiring ADHD to be reframed as a difference as opposed to a disorder, viewing of ADHD as a mismatch with their environments, and wanting the condition to be viewed through the lens of what it is like to embody the condition.

**Conclusions:** Incorporating patient-lived experiences into psychiatric diagnoses may increase diagnostic patient-centeredness and help healthcare providers better meet patient needs.

## Introduction

Attention-deficit/hyperactivity disorder (ADHD) is currently defined by symptoms of inattention, impulsivity, and hyperactivity (American Psychiatric Association, 2013). Although historically considered a disorder of childhood, ADHD persists into adulthood in 15-60% of children with ADHD (S. V. Faraone et al., 2006; Wender et al., 2001). Up to 90% of children with ADHD continue to have impairing symptoms into adulthood (Sibley et al., 2022). Adults with ADHD often disproportionately have unmet needs including substance use disorders (Capusan et al., 2019), interpersonal relationship concerns (Bruner et al., 2015; Cho & Park, 2016), incarceration (Baggio et al., 2018) and unemployment (Adamou et al., 2013; A. B. M. Fuermaier et al., 2021). Many children with ADHD lose access to services during transitions to adulthood, in part due to limited services and expertise surrounding adult ADHD (Swift et al., 2014). Although ADHD is highly treatable with stimulant medications (Stephen V. Faraone et al., 2021), the needs of people with ADHD could be better met by the medical system.

People with ADHD face stigma across the lifespan from peers, teachers, and family members, often resulting in internalization and self-stigma (Lebowitz, 2016; Nguyen & Hinshaw, 2020). Although comparatively few studies have explored ADHD-related physician stigma, physicians often have comparable levels of ADHD stigma compared with the lay population and endorse ADHD-related misconceptions (Anselm B. M. Fuermaier et al., 2012; Lebowitz, 2016; Moldavsky & Sayal, 2013).

Little research has investigated how adults with ADHD conceptualize their condition. Most children with ADHD regard the condition as a biological disorder, although some consider ADHD as part of their personality and others reject the diagnosis altogether (Wong et al., 2018). Qualitative studies investigating adults have shown that many view ADHD as a personality trait as opposed to a disorder (Lasky et al., 2016; Sedgwick et al., 2019; Waite & Tran, 2010) and regard ADHD as having positive aspects, such as promoting creative thinking and energy to pursue one’s passions (M. S. Brod et al., 2005; Hansson Halleröd et al., 2015; Schreuer & Dorot, 2017; Schrevel et al., 2016; Sedgwick et al., 2019). However, one study has reported heterogenous opinions from participants on whether they would like ADHD cured if possible (M. Brod et al., 2012). Understanding the perceptions that adults with ADHD have about their condition may help healthcare providers better meet this population’s needs.

This study aimed to use qualitative methods to characterize how young adults with ADHD conceptualize their condition and elicit suggestions they have for improving care for people with ADHD.

## Methods

### Ethical considerations

This investigation was carried out in accordance with the seventh revision of the Declaration of Helsinki. The following procedure was approved by the X [blinded for review] University Institutional Review Board (2000031327). Participants provided verbal consent before participation in the study.

### Procedure

Methods are described in full detail in a previous publication of data generated from the same focus groups (Ginapp et al., 2023); a brief description of methods is as follows. Participants were recruited from online sources including internet communities for people with ADHD such as Facebook and Reddit, with permission from the moderators of these spaces, and on the Children and Adults with ADD (CHADD) advocacy group website. This study was based out of the United States although participants from any country were eligible. Interested participants were first directed to a Qualtrics survey that screened for the inclusion criteria of being 18-35 years old, inclusively, reporting having been diagnosed with ADHD by a clinician, and scoring greater than or equal to 23 on the Adult ADHD Self-Report Scale (ASRS), reflecting on symptoms over the life course (Silverstein et al., 2019).

Participants were met individually before the focus groups to establish verbal informed consent. There were 146 completed responses to the screening survey, and 61 of these potential participants responded to follow-up emails. Of these, 51 potential participants completed informed consent for the study and 43 ultimately participated in focus groups. Reasons for not completing the study after responding were largely participant non-response without explanation, although some participants expressed that life circumstances had arisen that prevented them from continuing their involvement.

Twenty-seven (63%) of the participants had a confirmation of diagnosis by providing documentation of diagnosis, having their medical provider confirm diagnosis, or by the research team checking the State Prescription Monitoring program to see if they were being prescribed stimulants as a proxy for ADHD diagnosis. Participants who did not provide documentation were not excluded from the study. There were no instances of discrepancies between providers’ and participants’ reports of ADHD diagnoses, and all participants scored above threshold on the ASRS.

Nine focus groups over November and December 2021 were conducted via Zoom with three to six participants per group. Focus groups lasted approximately one hour and were video-and audio-recorded with participant consent. Focus groups were semi-structured and facilitated by a discussion guide which was comprised of questions asking 1) participants what they would like the medical community to better understand about ADHD and 2) if there were any misconceptions about ADHD they would like addressed. Participants were compensated with a $15 e-gift card, and after completion of all the focus groups, participants were provided with a summary document of the main themes.

### Theory and reflexivity

An interpretative phenomenological analysis (IPA) approach was employed to capture participant understanding of their lived experiences (Smith & Eatough, 2006). Although traditionally used for individual interviews, IPA has been adapted for use in focus groups with the unit of analysis remaining on the individual level (Love et al., 2020). We utilized focus groups to promote participant disclosure of potentially stigmatizing topics, as has been done in qualitative studies involving young adults with ADHD previously (Lefler et al., 2016).

The first author facilitated the focus groups, and the first and second author conducted the data analysis. The first author has been diagnosed with ADHD and is involved with online communities for those with ADHD; the second author has several family members with ADHD. This insight into the ADHD community allowed for knowledge about where to recruit participants, contributed to the development of the discussion guide, and provided context when analyzing the data. Potential biases include the researchers’ preconceived perceptions of ADHD based on their personal experiences and potential projection of these experiences onto participants. To mitigate these biases, the researchers journaled to reflect on their experiences and the possible biases they created and remained cognizant of these biases while conducting data analyses.

### Data analysis

Recordings were transcribed verbatim and uploaded to NVivo 12. Transcripts were first read independently and in their entirety by both researchers who performed data analyses and then read independently a second time to identify themes. The two researchers met to mutually develop a preliminary code book using inductive reasoning. Half of the transcripts were independently coded by the two reviewers using and modifying the preliminary codebook. Discrepancies were resolved by mutual agreement, and the code book was interactively updated. The remaining transcripts were coded with the updated code book by the lead researcher.

## Results

### Participant characteristics

Among the 43 participants, the median age was 29 years and median age of ADHD diagnosis was 22 years (range 5-34 years). The median time between diagnosis and entering this study was three years. Participants were 84% female, 72% White, 14% Asian, 9% Black, and 5% Hispanic/Latino. Eighty-six percent of participants were from North America, with the remaining being from Australia, Suriname, the Czech Republic, and the United Kingdom. Fifty-three percent were employed full-time, 44% were currently students, and 88% had at least some college education. The most common ADHD subtype was inattentive (42%), followed by combined (28%), and then hyperactive/impulsive (9%). The remaining participants either did not know their subtype or had been diagnosed before subtypes were introduced. Eighty-four percent of participants had been prescribed stimulant medication for ADHD, with nearly two-thirds reporting current stimulant use. Seventy-four percent of participants had been diagnosed with at least one other psychiatric condition, most commonly anxiety and depression. Sixty percent of participants were currently in psychotherapy for any reason. Most participants were recruited from Facebook (72%), followed by the Children and Adults with ADD (CHADD) advocacy group website (23%), and then Reddit (5%).

### Individual: the patient experience

#### Increased education and support

Participants expressed wanting to be educated on ADHD upon being diagnosed. Many felt that general practitioners in particular were unwilling to diagnose ADHD in adults, and those who did diagnose ADHD were often unable to provide support (beyond medications) such as psychotherapy or assistance with setting up accommodations. The perceived lack of support and education after having been diagnosed resulted in feelings of isolation in some participants.

> *I feel like when I was initially diagnosed it was very like, okay, you can’t do any school and you’re almost going to fail university, here’s max dosage Adderall, figure it out. And then not any kind of support with other people or really learning about what it’s like, so it felt like, okay, I know a bit more and I have some medication, but I still feel isolated in this experience*.
>
> *My general practitioner, I felt like this was way over their head, my diagnosis. They were able to give it to me and medicate me, but I feel like they had a checklist for me and that was it*.
>
> *I think that there needs to be more focus on these neurodevelopmental disorders, especially because they’re so common in the journey to become a doctor or psychologist, no, a doctor specifically, because a lot of the time your general practitioner is the first step to getting help, but they focus so little on it when it affects so many parts of your life*.
>
> Increased attention to ADHD was wanted in medical education to better equip providers to diagnose and manage the condition. This sentiment was especially expressed towards primary care education as primary care providers were often the first point of contact participants had with the medical system. Specialists were viewed as helpful, but having to access a specialist to receive medications or accommodations was viewed as burdensome.

#### Self-education and advocacy

Participants often took it upon themselves to learn about ADHD, in part because they felt they were not being provided adequate information about ADHD from their medical providers, and also because they anticipated that their providers would discredit them if they did not have research to back up their experiences. This resulted in participants feeling that they knew more about ADHD than their providers and were in fact educating them.

> *I do a lot of research, I felt like sometimes I was showing up more knowledgeable in the conversation, and I felt ashamed or even embarrassed to let on that’s how I felt*.
>
> *I walk into doctors ready to be told I’m wrong. No, I know my body, I know my mind. I do so much research and have so much prepared in case they tell me, “Oh no, you don’t have that.”*

Participants often put substantial effort into learning about ADHD. Self-advocacy was empowering to some participants, but more often resulted in feelings of shame when they felt they were educating their providers about ADHD.

#### Difficulty accessing stimulant medications

The stigma of people with ADHD being “drug-seeking” or abusing their stimulants was not directly covered in the discussion guide but came up independently in several focus groups. Hurdles participants had to overcome in order to access their medications included provider reluctance to prescribe and difficulties with insurance coverage. The desire to access stimulant medications was viewed as wanting their ADHD to be adequately treated; abuse of stimulant medications was regarded to mostly occur by those not diagnosed with ADHD.

> *It’s kind of crazy how much doctors withhold stimulants even though ADHD is one of the most treatable mental health disorders. If there’s anything I wish the medical field would recognize or know it’s that, stop being afraid to diagnose it and stop being afraid to treat it…. Do people abuse stimulants? Sure. Do people with ADHD abuse stimulants? No. People with diagnosed ADHD are not notoriously known for abusing stimulant medications*.
>
> *I would like a giant billboard for the medical community that says that very few of us are drug-seeking when we’re trying to get our stimulant medication, we just want to think and use our brain like everyone else. The fact that it’s so hard to get stimulants and the fact that psychiatrists in particular put up so many barriers to medication and insurance puts up so many barriers to medication, I feel like is some kind of human rights violation*.

Anger and frustration were widely shared in response to difficulties accessing stimulants, largely viewed as a result of stigma about prescribing to the adult population. It was important to participants to be viewed as people wanting to access efficacious medical treatment and not as drug-seeking.

### Structural: conceptualization of ADHD

“Different not disordered”

Participants wished for ADHD to be less pathologized by the medical community; viewed as a different way of thinking instead of a deficit of function. ADHD itself was viewed as having benefits and detriments, and participants wanted the condition to be framed without value judgments about “correct” ways of thinking.

> *[ADHD] is not a death sentence, it’s not a terminal illness, it’s not a disease. It’s a different perspective, it’s a different way of life almost. I mean if everyone had ADHD right, then the regular people who we see who aren’t diagnosed with ADHD perhaps we’d look at them and say they have hyperactive attention disorder, they have too much attention… It’s just a different mode I guess, a different layout. We just have different specs.*
>
> *I just want the whole mindset to change. It’s not an attention disorder, it’s not really a disorder. It is to people who don’t have an attention disorder, but to me who’s born with it and always grew up with it, it’s just a thing. It’s just the way I live, the way I work and function. Maybe it’s just me being in denial, but I just prefer the overall mindset changing to be more that it’s just a thing. It’s not necessarily a good thing or a bad thing, it’s just a thing*.

Participants challenged the current medical model which requires people to have an illness or disorder in order to access treatment. Many wanted an appreciation of different ways of thinking without a hierarchy of levels of functioning. ADHD was wanted to be framed as a difference that was neutral, not seen as better or worse than neurotypical ways of thinking.

#### Environmental problem

Some participants expressed that ADHD is not a disorder, but rather a byproduct of the way today’s society is structured. The educational system in particular was highlighted as an area that prioritizes sitting still and focusing on a certain task for long periods of time, and it might not be natural to function in this way.

> *Maybe it’s labeled as inappropriate because of for school, for example, it’s harder for teachers to get you to focus, but I feel like… rather than changing the people being molded by the school, we should change the mold, which is the school. We should accommodate more towards different ways of paying attention*.
>
> *This world’s not necessarily functioning in a way that I would want my brain to have adapted to. There’s something to be said for thinking differently and having a different way of functioning because I don’t necessarily feel the need to fit in exactly with the way the world is typically structured.*

Some participants expressed that they did not want to think in ways that are better adapted to the structure of today’s society. Instead of forcing people into a one-size-fits-all mold, multiple participants wanted more accommodations to different ways of thinking and learning.

#### Internal experience vs external presentation

Instead of ADHD being defined by external symptom manifestation, some participants wished that there was more consideration of the subjective experience of living with these symptoms. Symptoms of ADHD were noted to be framed negatively, as ways participants were bothersome to others (i.e., “does not seem to listen when spoken to directly”), instead of framing the symptoms as what it is like to embody them and in a more positive —or at least neutral— light (i.e., I find it helpful to have written instructions in addition to being told verbally). Other examples included “distracting others” instead of internal struggles to pay attention, or assigning value judgments to what is worthy of focus.

> *And I think there are probably better ways to frame that [ADHD] in way that captures the experience instead of it being a problem for somebody else*.
>
> *I think when they say it’s a deficit of attention, it’s a real judgement of what you should be paying attention to*.
>
> *Not a lot of people talk about it [ADHD] affects you, they just talk about how it affects other people. Like, “Oh, your hyperactivity, it’s just bothering all the other students at school.” Or, “Why can’t you pay attention, I’m trying to talk to you and it’s bothering me that you’re not listening.” And it’s like, “What about me? I’m trying to focus… I’m trying my best.”*

Participants felt that the framing of ADHD was more focused on how it was for others to interact with them as opposed to how they experienced their symptoms internally. This framing was viewed as oftentimes being judgmental or lacking empathy.

## Discussion

This qualitative study explores how young adults with ADHD wish ADHD was conceptualized by the medical community and how they believe the care they receive could be improved. Themes emerged around the individual experience of living with ADHD and accessing care and around considerations for how the medical community defines ADHD.

Study participants widely expressed that they wanted increased education about ADHD in the training of healthcare providers to improve competency for treating the condition, and for providers to be able to provide more comprehensive patient education. Information on ADHD diagnosis and treatments was seen as particularly lacking among general practitioners. Specialist care was seen as helpful but often difficult to access, especially without initial screening done by a primary care provider knowledgeable about ADHD. Given how common ADHD is in the adult population, clinician education on the condition should not be limited to psychiatrists.

Physician stigma about stimulant abuse was seen as a barrier to accessing medications by many participants. Rates of stimulant abuse among students are increasing, with 5-35% of college students reporting the non-medical use of stimulants (Clemow & Walker, 2014). An older review found that 5-22% of young adults abused their stimulant medications (Merkel & Kuchibhatla, 2009); however, stimulant abuse by individuals with ADHD has not been characterized in recent years. It is also unclear to what extent non-medical use of stimulants is from those with ADHD who are undiagnosed and are self-medicating. Although substance use disorders and ADHD often co-occur, treatment with stimulants has been shown to decrease risk of substance misuse among youth with ADHD (Dalsgaard et al., 2014; van Emmerik-van Oortmerssen et al., 2012). Efforts are needed to decrease stimulant misuse without decreasing access to treatment for those with ADHD.

Previous literature on how adults with ADHD think the condition could be differently conceptualized by the medical community has focused on viewing ADHD as a difference instead of a disorder. This viewpoint supports the neurodiversity movement which advocates that differences in neurodevelopment are natural variations and do not need to be pathologized (Leadbitter et al., 2021; Singer, 1998). The present findings of ADHD being a byproduct of not fitting into societal expectations, notably within the educational system, aligns with the social model of disability which frames disability not as biological in origin, but as a failure of societal accommodations for differences (Oliver, 1990). ADHD may also have beneficial features instead of being entirely a disability. Adults with ADHD have reflected that the condition may promote creativity and spontaneity and that learning to been cope with the struggles of ADHD can increase one’s growth, development, and awareness of oneself (Ginapp et al., 2022). Recent scholarship has remarked that the medical model of disability may benefit from incorporating the perspectives of the neurodiversity movement in order to provide better care to those affected by neurodevelopmental conditions (Sonuga-Barke & Thapar, 2021).

The findings that ADHD is currently defined by its external presentation and does not incorporate the subjective experiences of individuals living with the condition can be applied to psychiatric disorders more broadly. With the goal of increasing the reliability of psychiatric diagnoses, the use of objective operational criteria for psychiatric disorders was standardized starting with the DSM-III (Kawa & Giordano, 2012). Although there are benefits to formulating objective diagnostic criteria, the present findings indicate this approach may decrease the patient-centeredness of a diagnosis and oversimplify how physicians conceptualize mental health disorders.

ADHD-related stigma is widespread, both against ADHD symptoms and the diagnostic label (Lebowitz, 2016; Nguyen & Hinshaw, 2020). Findings from this study question to what extent stigma against the label of ADHD comes from within the medical community’s framing of ADHD as a disorder. Further research is necessary to explore this point and to evision how ADHD may be reconceptualized in a way that does not promote stigma and ableism.

This study has limitations. There was a high level of non-response among people who initially completed the screening survey, therefore making this a self-selecting group of people. There are likely perspectives not represented in this study as the sample was predominantly highly educated White women from North America who were diagnosed with the inattentive ADHD subtype in adulthood. It is possible that people with these demographics comprise the majority of members of the online communities from which participants were recruited, as the demographics of these communities are currently unknown. It is also possible that people with these demographics were more likely to participate in this study. It is hypothesized that because women and those with the inattentive sub-type may be more likely to go undiagnosed until adulthood, they may be more likely to seek out previously missing resources when they ultimately receive a diagnosis. As all participants in this study were engaged in online communities for those with ADHD, the perspectives of those with ADHD who are not connected with ADHD online communities may not be well represented here.

Although all participants self-reported a formal diagnosis of ADHD and scored above threshold on the ASRS, diagnosis was only confirmed in 63% of participants. Reasons for some participants not having a confirmed diagnosis largely consisted of participants verbally agreeing to have their healthcare provider contacted but then not providing electronic release-of-information documentation or the research team having been unable to establish contact with their provider. It is possible that some participants did not have a formal ADHD diagnosis and instead were self-diagnosed.

This qualitative study on experiences and perspectives of ADHD from young adults with the condition may shed light on how ADHD can be better treated and how the diagnosis of ADHD could be framed differently. The current conceptualization of ADHD, and psychopathology more generally, has been viewed by some as stigmatizing, and the medical system could likely benefit from increasingly considering perspectives from the social sciences and disability advocacy.

## Data Availability

All data produced in the present study are available upon reasonable request to the authors.

## Acknowledgements

We would like to thank the Facebook and Subreddit moderators and the Children and Adults with ADD (CHADD) advocacy group for their assistance with participant recruitment for this study. We would also like to acknowledge the QUA*Lab*, the Qualitative & Mixed Methods Lab, a collaboration between the Yale Child Study Center (New Haven, CT), and CESP, the Centre de recherche en Epidémiologie et Santé des Populations (Paris, France) for their insights in data analysis.

